# Neutralizing antibody titers predict protection from virus transmission in a cohort of household members with documented exposure to SARS-CoV-2

**DOI:** 10.1101/2024.12.10.24318774

**Authors:** Henrike Maaß, Imke Hinrichs, Martina Pavletic, Manuela Harries, Tatjana Prinke, Najat Bdeir, Richard Egelkamp, Berit Lange, Yannic C Bartsch, Mate Lerga, Luka Cicin-Sain

## Abstract

**Background:** While correlates of protection against symptomatic and severe breakthrough SARS-CoV-2 infections are well characterized, correlates of protection against virus transmission are incompletely understood.

**Methods:** We studied a Croatian cohort of individuals with documented household exposure to SARS-CoV-2 in December 2022. Sera were acquired prior to symptom onset, at the time of the COVID-19 diagnosis of the index cases, and comprehensively analyzed for correlates of protection against virus transmission. We monitored participants for 14 days and tested them with PCR at the end of the observation period to identify any virus transmission, including asymptomatic ones.

**Interpretation:** Out of nearly 200 tested serological parameters, 22 features were significantly different between the infected and the uninfected participants. Titers of variant-specific neutralizing antibody showed the biggest difference and were significantly higher in the uninfected subgroup. Some infected individuals with strong IgM responses to the spike antigen showed robust neutralization titers as well. Since IgM is likely an indication of recent antigenic exposure, data were reanalyzed by excluding such values. This refined analysis showed a complete segregation of infected and uninfected individuals into groups with low and high variant-specific neutralization titers. Therefore, our data indicate that high neutralizing titers are correlates of protection against SARS-CoV-2 transmission in intense contacts among household members.

**Funding:** This research was funded by the Impulse and Networking fund of the Helmholtz Association through the grant PIE-0008 to LCS and VH-NG-19-28 to YCB and by the Deutsche Forschungsgemeinschaft (DFG, German Research Foundation) under Germany’s Excellence Strategy - EXC 2155 - project number 390874280 to LCS. BL and MH received funding within the RESPINOW project from the Federal Ministry of Education and Research under the grant number 031L0298A.

**Research in Context:** *Evidence before this study:* Pre-existing immunity to SARS-CoV-2, whether from prior infections or vaccinations, has been shown to primarily protect against severe disease rather than preventing infection altogether. Many current studies examining this phenomenon focus on cohorts with breakthrough infections occurring a certain time after their last vaccination. However, these studies often lack precise information about when the individuals were infected and their serological status immediately before the infection.

*Added value of this study:* Unlike other studies, we focused on a cohort of individuals with a confirmed SARS-CoV-2-positive household member. Serum samples were collected before symptom onset, coinciding with the COVID-19 diagnosis of the index cases. We analyzed various serum features to comprehensively assess their ability to protect not only against severe disease but also against virus transmission. Our findings revealed that individuals who remained uninfected had significantly higher concentrations of neutralizing antibodies compared to those who became infected.

*Implications of the available evidence:* This finding suggests that neutralizing antibodies serve as a correlate of protection against virus transmission and could inform booster strategies based not on a fixed timeline but on antibody levels dropping below a specific threshold. However, due to the limited sample size of our study, larger studies are needed to confirm these results and establish an exact threshold.

## 1. Introduction

The coronavirus disease that emerged in December of 2019 (COVID-19), caused by the Severe Acute Respiratory Syndrome Coronavirus 2 (SARS-CoV-2), has had a profound impact on public health, leading to widespread infections, disease and death. The establishment of herd immunity due to the spread of infections and global vaccine campaigns has led to a reduction in severe symptoms. However, infection with SARS-CoV-2 can still result in long-term complications, known as long COVID, regardless of the initial severity of the disease (1), and vulnerable populations, such as the immunocompromised or the elderly, remain at risk of severe disease courses. Anti-COVID vaccines prevent severe disease, yet their effectiveness against symptomatic infections and milder disease courses wane relatively quickly (2). As a result, vaccination recommendations are primarily focused on individuals at higher risk of developing severe COVID-19, such as the elderly, immunocompromised individuals, and those working with vulnerable populations (3). Preventing infections in general, not just severe disease, should remain important in light of the risk of long COVID, which is common even upon less severe acute episodes, as it has been shown that vaccination reduces the risk of long COVID as well (4).

Since the development of SARS-CoV-2 vaccines, correlates of protection (CoP) have been studied to assess vaccine efficacy. Early research identified neutralizing antibodies (nAbs) as key CoPs against symptomatic disease induced by both vaccination and infection (5–7). However, no CoP or threshold levels have been established to guide recommendations on protection against infection, and hence, transmission. Therefore, national advisory bodies provide widely divergent guidelines on booster vaccination schedules. Most studies investigating CoPs have merely provided estimates based on the time since last vaccination, without knowing the exact time or circumstances of participants’ exposure to SARS-CoV-2 or objective criteria to quantify immune responses (8, 9). However, there are few studies assessing the role of serological parameters predicting the protection against virus transmission, especially in light of the rapidly evolving variants of SARS-CoV-2.

In this study, we examined households where one individual (the index case) tested positive for SARS-CoV-2 in the emergency department of the Clinical Hospital Center Rijeka, Croatia, during the winter of 2022/2023 to estimate the protection of transmission within the household using serological parameters and neutralizing antibodies. Household members were tested at an initial time point shortly after the index case and again 14 days later, or earlier if they developed COVID-like symptoms within two weeks. Based on their SARS-CoV-2 status during this period, household members were categorized into SARS2 positive and those that remained SARS2 negative throughout the analysis period and comprehensively analyzed to identify differences in serum parameters between those two groups. Variant specific nAbs, along with various other serum features were the best predictor of protection against virus transmission, while vaccination status and the timing of the last immunogenic event were non-significant as predictors of protection against breakthrough infections.

## 2. Material and Methods

### 2.1 Study participants and Study design

This study included household members of individuals who tested positive for SARS-CoV-2. Citizen who experienced SARS-CoV-2-like symptoms were tested in the Emergency department of the Clinical Hospital Center Rijeka. Household members (older than 18 years) of people who appeared SARS-CoV-2 positive in routine screenings were asked to visit the hospital within 48 h. Index (n=18) and household contacts (n=20) were tested via nasopharyngeal swab, by rapid test (Hangzhou AllTest Biotech Co., Ltd) as well as a fast RT-qPCR, essentially as described (10), to determine the SARS-CoV-2 status. Additionally, a blood sample was obtained. The household members who were SARS-CoV-2 negative at the first time-point (n=12 at 48 h) were asked to come back after 14-days or in between if they experience symptoms to get tested a second time. A schematic overview of the study design can be seen in Figure 1 and Figure S1. The recruitment of participants occurred in December 2022.

**Figure 1:**
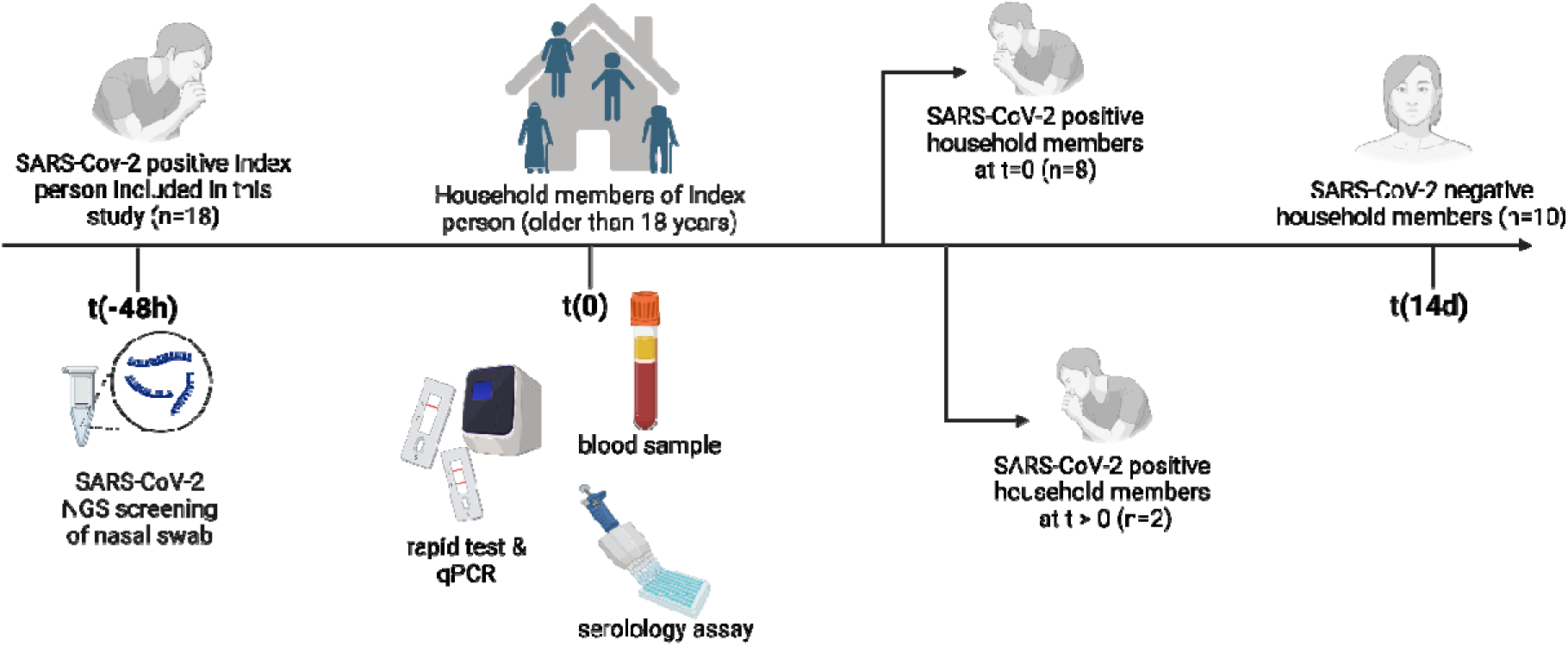
Brief schematic study overview. Household members of SARS-CoV-2 positive index persons donated a serum sample (t=0) at the Clinical Hospital Center Rijeka, 48 h after confirmation of the infection of the index person. Within 14 days, the household members were asked to report if COVID-19 like symptoms appeared or were asked to come back after two weeks to be tested by rapid test and qPCR. Serology features were determined usin various assays. Created with BioRender.com.

### 2.2 Sample preparation

Nasal swab samples were either stored at 4°C for immediate usage or aliquoted and stored at -80°C. The variant by which the index person was infected was identified by next generation sequencing (NGS) using the NEBNext ARTIC SARS-CoV-2 FS Library Prep Kit for Illumina with VarSkip 2b primers (New England Biolabs, MA, USA. Paired-end sequencing was performed on an Illumina NextSeq 1000 (Illumina, CA, USA). For assembly, CoVpipe v3.1.0 (Robert Koch-Institut, Germany; https://www.gitlab.com/RKIBioinformaticsPipelines/ncov_minipipe) was used with standard parameters, while lineage assignment was done with pangolin v4.3 and pangolin-data v1.21 (11). Whole blood samples were incubated for 30-60 min at room temperature. Serum and blood clot were separated by centrifugation at 1500 g for 10 min. The supernatant was aliquoted and stored at -80 °C. The samples were shipped on dry ice to the Helmholtz Center for Infection Research in Braunschweig for further analyses.

### 2.3 Demographic data and outcomes

Demographic data of all participants is listed in Table 1. Data was obtained through questionnaires, which were asked at the first and second sampling date either in person or through follow-up phone calls.

**Table 1:** Demographic data of household members included in this study, separated by the 14 day SARS-CoV-2 status.

|  | Household members (n=20) |  |  |
| --- | --- | --- | --- |
| Number of household members [median] | 2 (2-3) |  |  |
| COVID-19 qPCR Test | Positive (n=10) | Negative (n=10) | p |
| Female [%] | 20 | 30 | ns<sup>a</sup> |
| Number of Vaccinations [median, p25-75] | 2.5 (1.75-3) | 2 (2-3) | ns<sup>b</sup> |
| Days from latest vaccination until first test [median, p25-75] | 374 [341-475.5] | 409 [339-536.5] | ns<sup>b</sup> |
| prior COVID-19 infection | 4 | 7 | ns<sup>a</sup> |
| Days from latest immunogenic event until first test [median, p25-75] | 347 [338.5-387] | 312 [192.5-425.5] | ns<sup>b</sup> |
| Number of comorbidities | 1 [0-2.25] | 0.5 [0-1.25] | ns<sup>b</sup> |

<sup>a</sup> Calculated by Fisher's exact test. <sup>b</sup> Calculated by unpaired t test. ns = non-significant

### 2.4 Neutralization Assay

Neutralization assays were performed according to previously published protocol (12). The serum samples were heat inactivated at 56 °C and 500 rpm for 30 minutes and afterwards 1:10 diluted in DMEM (1% Penicillin-Streptomycin, 1% L-Glu, 5% FBS) and stored at 4 °C or -80 °C for long-term storage. Sera were two-fold diluted in a 96 well plate ranging from 1:100 to 1:102400 and incubated with pseudo- virus particles (600 pfu ± 30%) for one hour at 37°C. A control showing no inhibition by incubation in the absence of serum was used. Serum-virus samples were transferred to a VeroE6 96 well plate (80% confluent) and further incubated at 37 °C for 24 hours after which the plates were fixed with 4% paraformaldehyde (PFA) and kept at 4 °C until readout. To count GFP^+^ infected cells, the IncuCyte S3 (Sartorius) was used by performing whole-well scans (4x magnification) in phase contrast and green fluorescence settings (300 ms exposure). The IncuCyte GUI software (version 2019B Rev1 and 2021B) performed automated counting of GFP fluorescent foci. The pseudo-virus neutralization titer 50 (PVNT50) was calculated by non-linear regression model. The lower limit of confidence (LLOC) was set at a PVNT50 of 50. Non-responders were given a PVNT50 of 25 for visualization purposes.

#### 2.4.1 IgM depletion in serum samples

For certain experiments, only the IgG fraction of the nAbs was supposed to be measured. Therefore, IgM was depleted using beta-mercaptoethanol (2-ME) on heat-inactivated serum (13). Serum was incubated with 2-ME (final concentration of 140 mM) for 1h at room temperature and 300 rpm. Afterwards, the serum was diluted 1:10 with DMEM (1% Penicillin-Streptomycin, 1% L-Glu, 5% FBS) and immediately used for neutralization assays.

#### 2.4.2 SARS-CoV-2 specific IgM and IgG ELISA

To validate the efficiency of the IgM depletion by 2-ME, an enzyme-linked immunosorbent assay (ELISA) was performed. Therefore, serum was treated with 2-ME as explained before and the Human SARS-CoV-2 Spike (Trimer) IgM ELISA Kit (#BMS2324, Invitrogen, Waltham, United States) and Human SARS-CoV-2 Spike (Trimer) IgG ELISA Kit (#BMS2325, Invitrogen, Waltham, United States) were performed as per manufacturer’s instructions. The assay was run on exemplary participants with high, medium and low IgM based on the Luminex assay, in technical duplicates. The Infinite® F50 (Tecan, Männedorf, Switzerland) was used for readout of the ELISA plates.

### 2.5 Detection of antigen-specific antibody isotypes and Fc-receptor binding

Human serum samples were profiled for antigen-specific antibody isotypes and binding of Fcγ receptors with a custom antigen panel as previously described (14). Recombinant antigens were obtained commercially (Sino Biological, Beijing, China). The custom antigen panel consisted of recombinant SARS-CoV-2 trimeric spikes (D614G, B1.1.529 and BA.4/BA.5/BA.5.2), receptor binding domain (RBD) (D614G, B1.1.529 and BA.4/BA.5/BA.5.2), N-terminal domain (NTD) (2019-nCoV), nucleocapsid (NC) (2019-nCoV), S1 (D614G) and S2 domains (2019-nCoV), the trimeric spike of the human coronaviruses HKU1 (N5) and OC43, and the respiratory syncytial virus (RSV) prefusion protein (preF). For the Luminex assay, 10 µg antigen were covalently coupled to 2 million carboxylated magnetic microspheres (Diasorin, Italy) by carboxy-coupling as described before (14).

Serum samples were diluted with assay buffer (PBS, 0.1% BSA) depending on detector to 1:500 for IgG1, 1:100 for IgG2-4, IgA1-2 and IgM, and 1:1000 for the Fcγ receptors. In a 384-well plate, 5 µL of diluted sample were incubated overnight with a pool of the antigen-coupled microspheres (1,000 beads per antigen per well) at 4 °C. Plates were washed three times with PBS-T (PBS, 0.05% Tween-20) and incubated for 1 h with 40 µL of PE-conjugated secondary detector (1 µg/ml) against IgG1 (RRID: AB_2796628), IgG2 (RRID: AB_2796639), IgG3 (RRID: AB_2796701), IgG4 (RRID: AB_2796693), IgM (RRID: AB_2796577), IgA1 (RRID: AB_2796656) and IgA2 (RRID: AB_2796664) (all Southern Biotech, AL, USA). To detect Fc receptor interaction, streptavidin-PE:biotin-coupled FcγRIIa (H167), FcγRIIa (R167), FcγRIIb/c, FcγRIIIa (F176), FcγRIIIa (V176), FcγRIIIb (NA1) and FcγRIIIb (NA1) (all AcroBiosystems, DE, USA) were used (1 µg/mL). Upon incubation, immune complexes were washed three times with PBS-T using the HydroSpeed automated plate washer (Tecan, Switzerland), resuspended in assay buffer and acquired at the ID7000 Spectral Cell Analyzer (Sony Biotechnology, CA, USA).

### 2.6 Biotinylation and bead-coupling of antigens

For antibody-dependent complement deposition (ADCD), antibody-dependent THP-1 cell-phagocytosis (ADCP) and antibody-dependent neutrophil-phagocytosis (ADNP), the spike trimers of wild-type SARS- CoV2 (D614G) and Omicron variant BA.4/5 were biotinylated using the NHS-Sulfo-LC-LC kit (Thermo Fisher, MA, USA) according to the manufacturer’s instructions. After the reaction, excess biotin was removed by size exclusion using Zeba spin desalting columns with a 7 kDa cut-off (Thermo Fisher). 10 µg of biotinylated antigen was coupled to 10 µl of 1.0 µm NeutrAvidin-labelled microspheres for 2 h at 37 °C.

### 2.7 Antibody-dependent complement deposition (ADCD)

The ability of antigen-specific antibodies in the human serum samples to induce the complement cascade was assessed as described before (15). Briefly, immune complexes were formed in a 96-well round bottom plate by incubating antigen-coated microspheres with 10 µL of 1:10 diluted human serum samples for 2 h at 37 °C. Reconstituted guinea-pig complement (Cedarlane, ON, Canada) was cleared by centrifugation, transferred to GVB++ buffer (Novateinbio, MA, USA) and incubated with washed immune complexes for 20 min at 37 °C and 5% CO_2_. Complexes were then washed once with 15 mM EDTA in PBS. Deposited C3 on beads was stained with 1:100 diluted anti-guinea pig C3a-FITC antibody (MP Biomedicals, CA, USA) and afterwards fixed with 4% paraformaldehyde (PFA). C3 deposition was then determined as mean fluorescent intensity (MFI) on an ID7000 Spectral Cell Analyzer (Sony Biotechnology).

### 2.8 Antibody-dependent neutrophil-phagocytosis (ADNP)

Neutrophil-mediated phagocytosis of serum antibodies was determined as previously described (16). Antigen-coated yellow-green fluorescent microspheres were incubated with 10 µL of 1:50 diluted human serum for 2 h at 37 °C and 5% CO2. Primary neutrophils were obtained by lysis of whole blood with Ammonium-Chloride-Potassium (ACK) buffer (Biomol, Germany) and transferred to R10 media (RPMI 1640, 10% FCS, 1x GlutaMax, 1x Penicillin-Streptomycin, 10 mM HEPES). The immune complexes were washed by centrifugation and incubated with 50,000 neutrophils for 1 h at 37 °C and 5% CO_2_. Next, neutrophils were stained with 1:100 diluted anti-human CD66b-Pacific Blue antibody (BioLegend, CA, USA, RRID: AB_2563294) and fixed with 4% PFA for 20 min. Phagocytic activity of neutrophils was measured on an ID7000 Spectral Cell Analyzer (Sony Biotechnology). A phagocytosis score was calculated as product of frequency bead positive neutrophils and the MFI of bead positive neutrophils.

### 2.9 Antibody-dependent THP-1 cell-phagocytosis (ADCP)

The level of serum antibody-induced phagocytic activity of THP-1 monocytes was detected as previously described (5). In brief, antigen-coated yellow-green fluorescent microspheres were combined with 10 µL of 1:100 diluted human serum in a 96-well round bottom plate and incubated for 2 h at 37 °C and 5% CO2. Next, 25,000 cultured THP-1 cells (#ACC 16, DSMZ, Germany) in R10 media were added and incubated overnight at 37 °C and 5% CO_2_. THP-1 cells were then fixed with 4% PFA for 20 min, washed and resuspended in PBS prior to acquisition on an ID7000 Spectral Cell Analyzer (Sony Biotechnology). A phagocytosis score was calculated for THP-1 monocytes in the same way as for neutrophils.

### 2.10 Antibody-dependent NK cell-activation (ADNKA)

To investigate antibody-mediated NK cell activation, 96-well flat bottom high-binding plates were coated with 50 µL (3 µg/mL) of either the spike trimer of wild-type SARS-CoV2 (D614G) or Omicron variant BA.4/5. After blocking with 5% BSA-PBS for 2 h, 50 µL of 1:30 diluted human serum samples were added and incubated overnight at 4 °C. Primary NK cells were derived from leukocyte reduction cones of healthy donors using the RosetteSep NK cell enrichment kit (STEMCELL Technologies, MA, USA) according to the manufacturer’s instructions followed by density gradient centrifugation. The enriched cells were then stimulated with 1 ng/mL of recombinant human IL-15 (BioLegend) overnight at 37 °C and 5% CO2. After washing the immune complexes three times, 50,000 NK cells were added to each well and incubated with 2 µg Brefeldin A (Sigma Aldrich, MA, USA), 1:80 GolgiStop (BD Biosciences, NJ, USA) and 1:11 diluted anti-human CD107a-BV605 (BioLegend, RRID: AB_2563851) for 5 h at 37 °C and 5% CO2. To define NK cells as CD3-CD56+, the cells were stained for 15 min with 1:40 diluted anti-human CD3-APC-Cy7 (BioLegend, RRID: AB_830755) and 1:20 diluted anti-human CD56-PE-Cy7 (BioLegend, RRID: AB_2857328). After fixation and permeabilization with fixation medium (Invitrogen) and permeabilization medium (Invitrogen), respectively, NK cells were stained intracellularly with 1:200 anti-human MIP-1β-BV421 (BD Biosciences, RRID: AB_2737877) and 1:50 anti-human IFN-γ-PE (BioLegend, RRID: AB_315440) for 15 min. Finally, samples were washed, resuspended in PBS and the MFI of extracellular and intracellular markers acquired on an ID7000 Spectral Cell Analyzer (Sony Biotechnology).

### 2.11 Statistical analysis

Due to small sample size, all data was log10-transformed and analyzed using parametric tests. For group comparison, an unpaired t-test with Welch’s correction was performed. Data representation and statistical analysis was done using R software v4.3.2 or GraphPad Prism v10.2.2 (GraphPad Software, CA, USA).

### 2.12 Ethical Considerations

The Institutional Review Board of the Rijeka Clinical Hospital Center (2170-29-02/1-22-2) as well as the Hanover Medical School (10783_BO_K_2023) approved this study. Written informed consent was obtained from all participants.

## 3. Results

### 3.1 Demographic and clinical characteristics of study population

This study was conducted at the Clinical Hospital Center Rijeka, focusing on households in which on member tested positive for SARS-CoV-2 (from now on called index person). Household member donated a blood sample within 48 h after the index person was tested. The samples were analyzed, categorizing the household members based on whether they contracted the infection within 14 days after the initial sampling or remained uninfected. A schematic study overview can be seen in Figure 1 and Figure S1.

The study cohort consisted of 18 index people and 20 household members. Half of the household members (n=10) remained negative over the 14 day study period (from now on called not-infected or negative group). Eight household members were SARS-CoV-2 positive at the initial testing and two wer infected within the 14 days (these 10 participants will be referred to as infected or positive group). Both groups presented similar characteristics, which can be seen in Table 1. In summary, both groups showed no significant differences in terms of number of vaccinations or days since last vaccination or latest immunogenic event (i.e., vaccination or natural infection). The non-infected group presented non-significantly higher number of participants with a previous COVID disease.

### 3.2 Not-infected household members show multiple differently expressed features of adaptive humoral immunity

We quantified various features of spike-specific humoral immunity in the initial serum samples, including a systems serology approach, levels of antibodies targeting different virus epitopes as well as different SARS-CoV-2 variants and respiratory viruses like RSV (Table S1). Additionally, nAbs targeting different SARS-CoV-2 spike strains were analyzed. Out of 198 features, 21 were significantly upregulated in the negative group, compared to the positive and one downregulated in the negative group (Figure 2a). Amongst those, seven features were IgG1 binding titers, targeting different SARS-CoV-2 epitopes or strains and four were related to binding of IgA1 antibodies. Additionally, the nAb concentrations against all tested SARS-CoV-2 strains were increased in the not-infected group. Figure 2b shows the log-transformed levels of the 22 differently expressed features.

**Figure 2.**
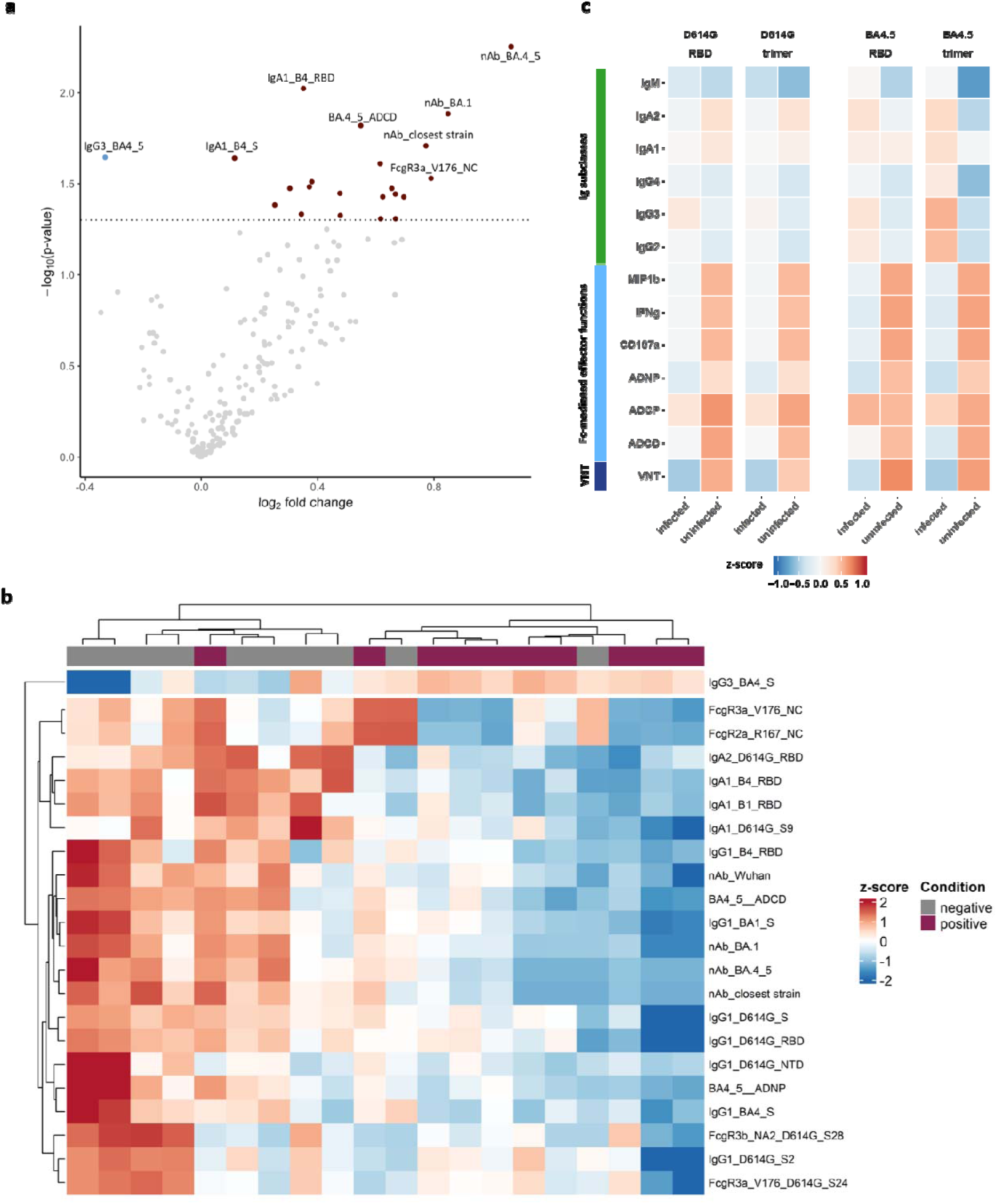
Differentially expressed features in SARS-CoV-2 infected household members at initial sampling. a) This figure displays a volcano plot of differentially expressed features in household members who were or becam SARS-CoV-2 positive within 14 days after initial testing. The dotted line marks a p-value threshold of 0.05. Gre dots represent non-significant features. Blue dots indicate features significantly upregulated in positive househol members, while red dots represent features significantly upregulated in negative household members. b) A heatma illustrating the significantly differentially expressed features identified in the volcano plot. Feature levels were log-transformed. Each column corresponds to a participant, and each row represents feature levels. Column annotations denote the condition of each participant. Clustering was performed based on Euclidean distance. c) Heatmaps of normalized average IgG1 corrected values (feature/IgG1) for each feature are shown for spike trimers and RBD of WT (D614G) and Omicron BA.4.5, comparing infected (n=10) to non-infected (n=10) individuals. Data from Luminex bead assays, ADCD and virus neutralization assays were log10 transformed prior to IgG1 normalization.

As some participants presented high IgG1 levels, which is usually the most dominant IgG subclass, we normalized the data by the IgG1 signal of uninfected and infected groups to see whether there are mechanistic differences in FcgR binding, Fc function or neutralization (Figure 2c). We identified Fc- mediated effector functions and the virus neutralizing titers (VNT) to be more robust in uninfected participants, which is in line with the findings of Figure 2b and c, and highlights the importance of nAb as CoP.

### 3.3 nAb as correlate of protection

To further investigate the importance of nAbs, we individually analyzed their concentrations. All concentrations were elevated in participants who did not get infected (Figure 3a) even against the original Wuhan strain, which by the time of sampling was not present anymore. Interestingly, the closer the tested strains got to the strain by which the index person was infected and to the at that time circulation variant BA.4/5, the more prominent the difference appeared. By looking at their prognostic potential with receiver operation characteristic (ROC) curve, all measured nAb concentrations showed significance (Wuhan p = 0.028; BA.1 p=0.0191; BA.4/5 p=0.0156; closest strain (c.s.) p=0.0091) (Figure 3b).

**Figure 3.**
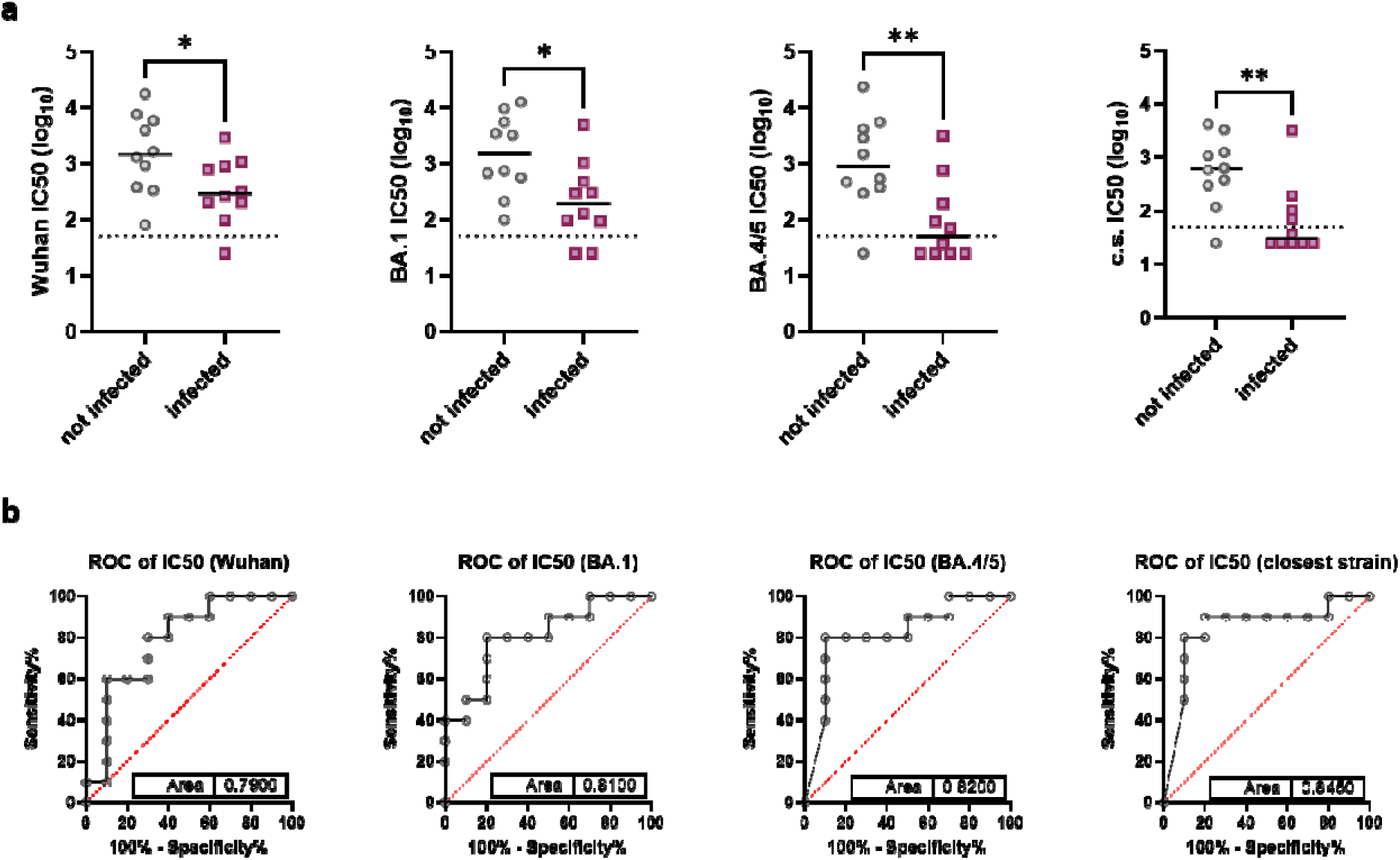
Neutralizing antibody concentrations in household members at initial sampling. a) Neutralizin antibody (nAb) concentrations were measured using pseudovirus neutralization assays against SARS-CoV-2 spik variants Wuhan, Omicron BA.1, and BA.4/5, as well as a relatively closely related strain. Participants were categorized as not infected (grey dots) or infected (purple squares). The dotted line indicates the limit of confidence. For analysis, the data was log-transformed and a t-test with Welch’s correction was conducted. The black line represents the median for each group. b) ROC analysis of nAb concentrations, with the area under the curve shown in the plot. * = p < 0.05.

### 3.4 IgM-high participants screw protection potential of nAb

As the majority of our infected participants were already SARS-CoV-2 positive at the initial time point, we expected high IgM levels in some of them, limiting our ability to mainly analyze the nAb fraction which was present before the exposure. Hence, we checked the IgM levels targeting the nucleocapsid of the household members to identify outliers with very high IgM. In Figure 4 we show the levels being not significantly different between the two groups. Nevertheless, we identified three outliers (one in the non-infected, two in the infected) which had extremely elevated IgM in contrast to the rest of their respective group. We compared the neutralizing activity after excluding these participants with outlier IgM levels as they might have had a previous infection and their results would not be in line with the other participants. The significance of all variants increased drastically, separating the concentration of both groups clearly (Figure 4b). Moreover, the ROC analysis emphasize what impact those outliers have on the predictability of this feature (Wuhan p=0.0021; BA.1 p=0.0015; BA.4/5 p=0.0005; c.s. p=0.0008) (Figure 4c).

**Figure 4.**
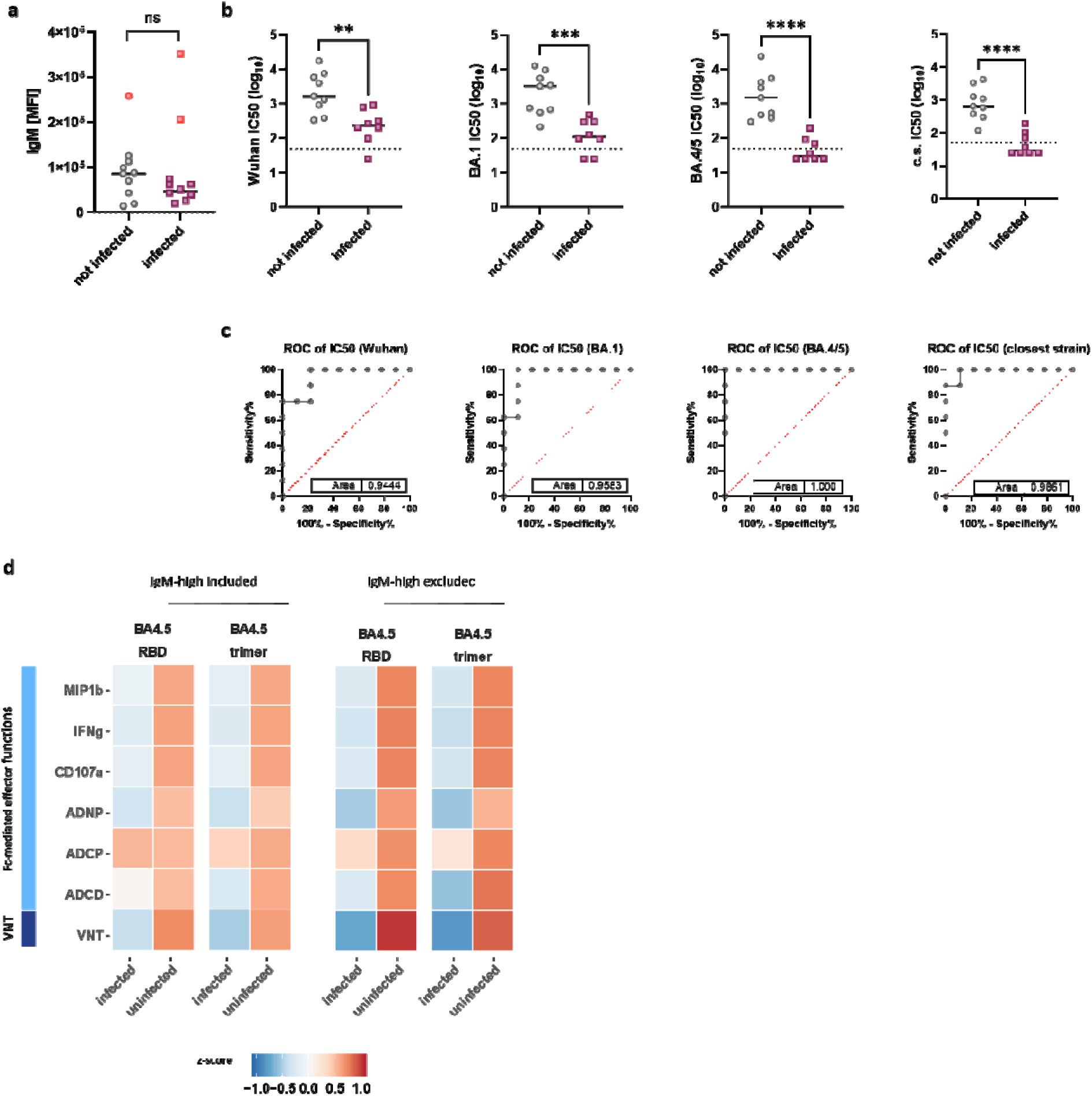
nAb concentration wihtout IgM outliers. a) IgM levels against the SARS-CoV-2 nucleocapsid were measured using Luminex. Participants were categorized as not infected (grey dots) or infected (purple squares). One particpant in the not infected group (red dot) and two outlier in the infected group (red squares) presented a high IgM level. A t-test with Welch’s correction was conducted for analysis. b) Participants were categorized as not infected (grey dots) or infected (purple squares), participants with high IgM were excluded for the analysis. The dotted line indicates the limit of confidence. For analysis, the data was log-transformed, and a t-test with Welch’s correction was conducted. The black line represents the median for each group. c) ROC analysis of nAb concentrations, with the area under the curve shown in the plot. d) Heatmaps of normalized average IgG1 correcte values (feature/IgG1) for each feature are shown for spike trimers and RBD of WT (D614G) and Omicron BA.4.5, comparing normalization including IgM-high participants (left) to exluding IgM-high participants (right). Data from Luminex bead assays, ADCD and virus neutralization assays were log10 transformed prior to IgG1 normalization. MFI = median fluorescent intensity; * = p < 0.05; ** = p < 0.01; *** = p < 0.001.

Additionally, by using 2-ME, we were able to eliminate IgM out of the serum (13). We validated the results by ELISA (Figure S2a) confirming that IgM was depleted with modest effect on IgG concentrations. Next, all pseudovirus neutralization assays were repeated with 2-ME treated serum (Figure S2b). However, the difference between the two groups did not drastically increase and in the case of the closest strain even decreased (ROC Wuhan p=0.0284; BA.1 p=0.0233; BA.4/5 p=0.0156; c.s. p=0.0233). Interestingly, individuals with initial los neutralizing captivity showed increased neutralization upon IgM depletions.

Lastly, we repeated the normalization by IgG1 as shown before but excluded the three IgM high participants, and saw even more prominent Fc-mediated effector functions and VNT responses in uninfected participants (Figure 4d). This summarizes the overall finding of nAbs being one of the most important CoP.

## 4. Discussion

SARS-CoV-2 vaccination has been suggested to primarily protect against symptomatic or severe disease rather than preventing infection altogether (17). Consequently, vaccinations are mainly recommended for individuals at high risk for severe COVID-19, such as those over 60 years old, immunocompromised individuals, or healthcare workers interacting with high-risk groups (3). Previous studies with a similar aim, but no exposure documentation, have highlighted the importance of nAbs in disease prevention (18–20). This study aimed to determine whether any serological parameters of acquired immunity may predict protection from breakthrough infections. A previous study showed that healthcare workers with breakthrough SARS-CoV-2 infections has significantly lower IgG antibody titers, but showed substantial overlaps between groups (21). However, they did not document the exposure of study participants to the virus, it remained unclear if participants who remained negative were protected by their adaptive immune response or merely unexposed to the virus. Another study that was conducted in Israel in 2021 tested people exposed to a SARS-CoV-2-positive contact in their household during the Delta wave. This study identified nAb and IgG titers as CoPs against breakthrough infection, with some outliers that were infected even when antibody titers were rather high. However, that study tested only antibodies against the ancestral spike antigen, and not the Delta variants, which was in circulation at the time (22).We conducted our study with households containing a SARS-CoV-2 positive index person, who acted as the disease transmitter and assessed serology in a variant specific manner. Household members provided blood samples within 24 hours and were monitored for infection and symptoms for 14 days thereafter, upon which they donated a blood sample again. Serum features of targeted responses to ancestral, Omicron BA.1 or BA.5 spike antigen were performed, including nAbs.

The analysis of approximately 200 features in the serum samples from the early time point revealed 22 significantly differentially expressed markers in our study. Half of these markers were related to Ig subtypes, as for instance IgG1 and IgA, or nAbs titers. A similar study conducted in Israel with 1,461 household members in 2021, when the Delta variant was dominant, found results consistent with ours, identifying IgG titers and nAbs as correlates of protection against infection and disease severity (22). However, that study did not comprehensively address parameters of antibody responses in a virus variant specific manner. Hence, the ability of antibodies to neutralize the Delta variant were not assessed. Our study shows that neutralization assays against the ancestral variant are less accurate in predicting the protection against infection than those based on antigens from contemporary circulating virus variants. While antibodies against pre-Alpha spike antigen were still significantly upregulated in individuals who remained uninfected, the ROC analysis revealed that their predictive accuracy was inferior to those of closely related variants.

Importantly, both groups in our study were similar in terms of vaccination status or time elapsed since the last immunogenic event. This suggests that the time since the last vaccination is a rather crude predictor of protection against breakthrough infections. On the other hand, neutralizing titers against a close variant were a very robust correlate of protection, especially among people with no immunological evidence of a recent infection. We propose that our results argue for two important ideas. Firstly, our data argue that high neutralizing titers may protect against any breakthrough infection, and thus impede the transmission. Therefore, a high antibody titer induced by a fresh booster shot would prevent not only disease, but also break the chain of transmission. Secondly, our data provide evidence that eliciting high neutralizing titers among caregivers of immunocompromised people would limit the spread of the virus to the vulnerable and thus protect the population at risk. Consequently, we propose that regular assessments of neutralizing antibody titers of medical personnel and caregivers working in senior foster care would identify those who need additional booster shots to protect indirectly the very vulnerable population, whose immune system is weak and frequently subject to vaccine failure.

Since some participants were already infected at the initial time point, we investigated the impact of IgM on neutralization. We observed that IgM levels were elevated in several study participants, indicating recent exposure to the virus. Excluding the participants with high IgM levels from the analysis revealed a remarkably strong differentiation in nAb concentrations between positive and negative household members. This suggests that individuals with high IgM levels may skew the predictive value of nAbs as a correlate of protection. To explore this further, we also depleted IgM using 2-ME and repeated the neutralization assay (13). Overall, no significant differences were observed compared to the IgM-containing assays. This finding aligns with the results of Klingler *et al*., who demonstrated that IgM and IgG are both important for virus neutralization (23). We propose that the participants with high IgM had high neutralization efficacy due to recent infections and that any serological diagnostics for CoP needs to include IgM testing to identify people with a recent exposure to the virus.

Our study has several limitations. We performed exhaustive immunological analyses on samples from a rather small cohort. In consequence, the statistical power was low and some of the features that were classified as non-significant may well differ between people who are protected against a household transmission and those who are not. The low statistical power also limits our ability to define quantitative thresholds in antibody titers needed for protection. Moreover, since we used a neutralization assay performed with pseudotyped viruses, the standardization of this assay for quantitative purposes may present technical challenges. We did not interview the household member about their behavior, such as isolation from the index case, and we did not perform PCR analyses on a daily basis, thus potentially omitting cases that resolved very rapidly. Additionally, our study was conducted in late 2022, during the prevalence of the Omicron BA.4/5 variant, prior to the emergence of currently circulating variants. Hence, a similar study performed on samples from current and upcoming variants is required to solidify our conclusions.

Nevertheless, our findings demonstrate that nAb concentrations strongly correlate with protection against SARS-CoV-2 infection. Contrary to current vaccination recommendations, we propose that serology testing for nAbs and boosting those with low concentrations may prevent not only disease manifestations, but also effectively block transmission. Notably, testing with the variant that is currently prevalent provides high accuracy, eliminating the need for an index person-specific test and simplifying this diagnostic approach.

## Supporting information

Serolomics data for all study participants

## Data Availability

All data produced in the present work are contained in the manuscript

## Acknowledgement

We gratefully acknowledge Denise Clesle, Ayse Barut and Yuliia Polianska for technical support, Natascha Gödecke for administrative support with regulatory affairs and compliance and Henning Jacobsen for technical advice and discussions. IH was supported by the Hannover Biomedical Research School (HBRS) and the Center for Infection Biology (ZIB), Germany.

## Author contribution

Conceptualization: H.M., M.L., M.H., B.L., M.P. and L.C.-S.; investigation: H.M., I.H., M.L., T.P., N.B. and R.E.; formal analysis: H.M., I.H. and L.C.-S.; resources: M.L. and M.P.; data curation: H.M. and L.C.-S.; writing – original draft preparation: H.M.; writing – review and editing: L.C.-S.; supervision: M.L. and L.C.-S.; funding acquisition: Y.B. and L.C.-S. All authors have read and agreed to the published version of the manuscript.

## Data availability

The dataset used is available in the supplementary file and from the corresponding author on reasonable request.

