## Supplementary figures 1-3 for "Neutralizing antibody titers predict protection from virus transmission in a cohort of household members with documented exposure to SARS-CoV-2"

### **Title**

### **Contents**

**Table S 1**: Biomarker assays

**Figure S 1**: Detailed schematic study overview

**Figure S 2**: Closest strain distribution

**Figure S 3**: IgM depletion by beta-mercaptoethanol

**Table S 1: Biomarker assays**. For each antigen (left) Ig-subclasses and FcgR (right) were determined by Limunex, totaling in 182 biomarkers.

| **Antigen** | **Detector** |
| --- | --- |
| D614G S1+S2 trimer | IgG1 |
| D614G S1 | IgG2 |
| D614G RBD | IgG3 |
| 2019-nCoV S2 | IgG4 |
| 2019-nCoV NTD | IgA1 |
| 2019-nCoV Nucleocapsid | IgA2 |
| BA.1.1.529 S1+S2 trimer | IgM |
| BA.1.1.529 RBD | FcgR2a (H167) |
| BA.4/BA.5/BA.5.2 S1+S2 trimer | FcgR2a (R167) |
| BA.4/BA.5/BA.5.2 RBD | FcgR2b/c |
| OC43 S protein | FcgR3a (F176) |
| HKU1 S protein | FcgR3a (V176) |
| RSV F protein | FcgR3b (NA1) |
|  | FcgR3b (NA2) |

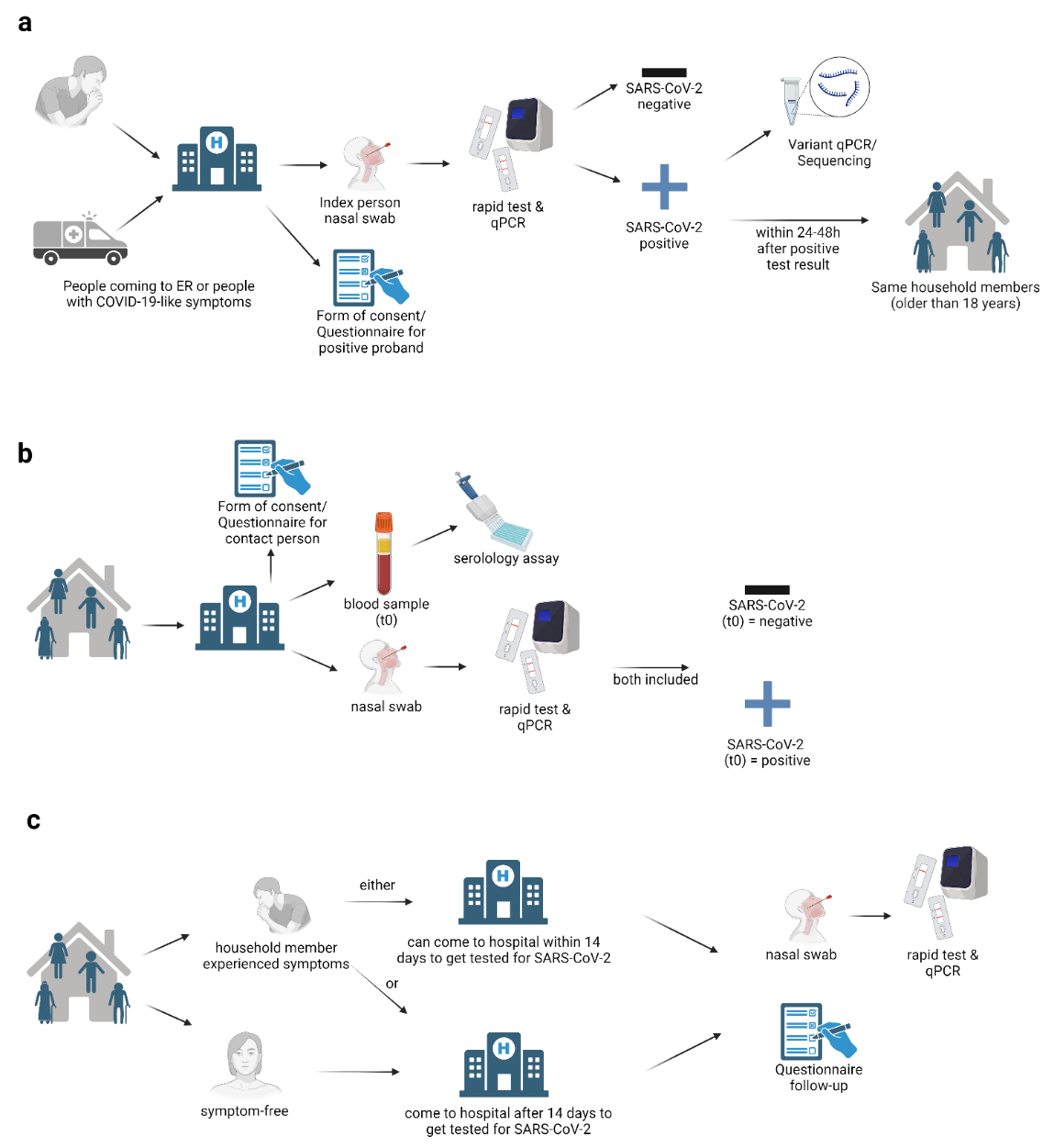

**Figure S 1: Detailed schematic study overview**. a) Citizens admitted to the emergency department (ER) of the Clinical Hospital Center Rijeka or those who came for regular COVID-19 testing were tested for SARS-CoV-2 using a rapid test and qPCR. If the citizen tested positive, the variant was identified via NGS, and their household members were asked to visit the ER within the next 48 hours. b) Household members were requested to fill out a questionnaire and donate a blood sample. They also underwent a rapid test and qPCR to determine their SARS-CoV-2 status at the initial time point (t0). c) Within 14 days, participants who experienced COVID-like symptoms were asked to return to the ER for retesting. Symptom-free participants returned after 14 days for repeat testing with a rapid test and qPCR, and another blood sample (t1) was collected. Created with BioRender.com.

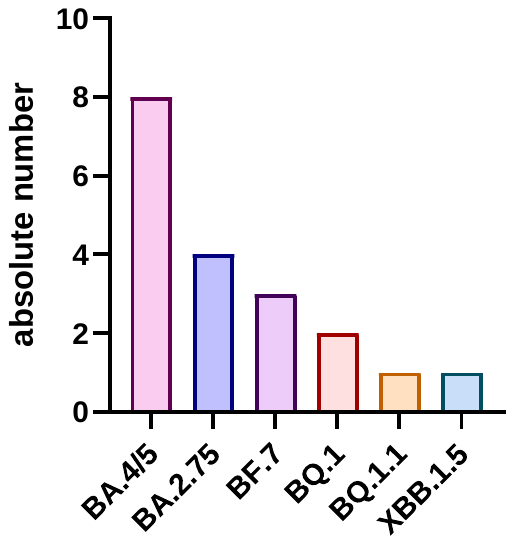

**Figure S 2:** **Closest strain distribution.** SARS-CoV-2 variants were determined by NGS and a closely related strain was identified. Pseudovirus neutralization assays were run with a virus expressing the relative spike variant.

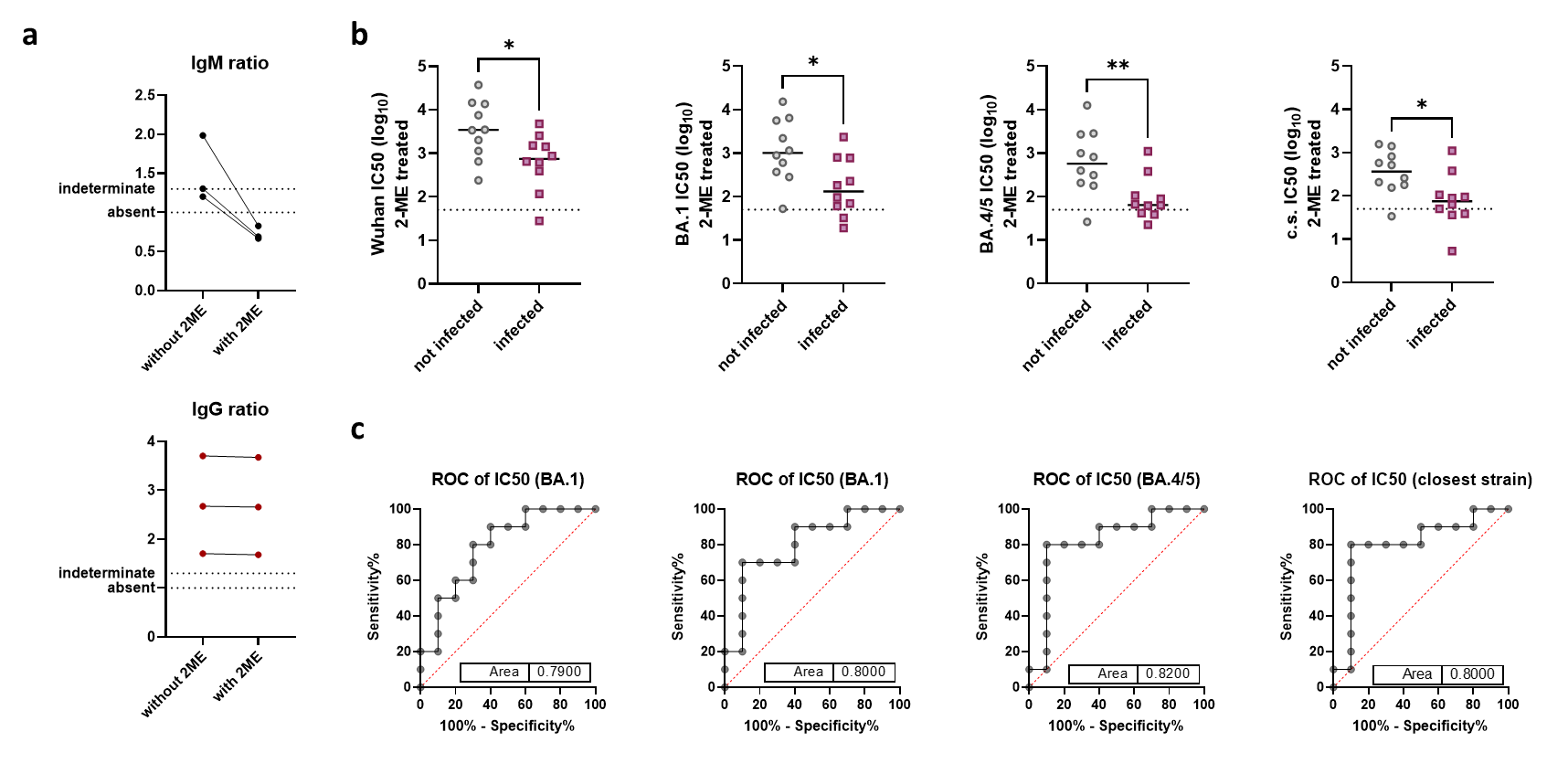

**Figure S 3:**  **IgM depletion by beta-mercaptoethanol**. a) samples of all participants were pre-treated with 2-ME for 1h. The efficiency of 2-ME treatment was validated by ELISA using exemplary samples. IgM (top, black dots) and IgG (bottom, red dots) concentrations are shown without and with 2-ME treatment. b) Ppseudovirus neutralization assays were performed to detect nAbs against SARS-CoV-2 spike variants Wuhan, Omicron BA.1, and BA.4/5, as well as a relatively closely related strain. Participants were categorized as not infected (grey dots) or infected (purple squares). The dotted line indicates the limit of confidence. For analysis, the data was log-transformed, and a t-test with Welch’s correction was conducted. The black line represents the median for each group. c) ROC analysis of nAb concentrations, with the area under the curve shown in the plot. * = p < 0.05; ** = p < 0.01.
